# The indirect effect of the bivalent human papillomavirus vaccination program: an observational cohort study

**DOI:** 10.1101/2025.02.28.25323074

**Authors:** Marit Middeldorp, Janneke W. Duijster, Mirjam J. Knol, Birgit H.B. van Benthem, Johannes Berkhof, Audrey J. King, Hester E. de Melker

## Abstract

**Background:** The impact of human papillomavirus (HPV) vaccination programs depends on the degree of indirect protection against new infections achieved among unvaccinated women. We estimated the indirect effect of bivalent HPV vaccination by comparing the HPV-type incidence in unvaccinated female participants between a cohort offered vaccination in 2009/10 and a cohort of similar aged women offered vaccination in 2014.

**Methods:** We compared the incidence rates of HPV types in the HAVANA cohort (follow-up from 2010/11 until 2015/16) with those from the HAVANA-2 cohort (2017-2022) using two regression approaches to estimate the indirect effect of HPV vaccination. First, we calculated the incidence ratio (IRR) for a vaccine or cross-protective type in HAVANA-2 versus HAVANA by Poisson regression and compared it to the IRR for a non-cross-protective type. The indirect vaccine effect is defined as 1-ratio of the IRRs. Second, we performed Cox regression with infection by vaccine or cross-protective type as the endpoint and calculated the hazard ratio (HR) for HAVANA-2 versus HAVANA after adjusting for time-varying sociodemographic variables. The indirect effect is defined as 1-HR.

**Results:** We included 661 unvaccinated participants in HAVANA and 927 in HAVANA-2. We observed a significant reduction in incident HPV16 infections of 70.9% (95% CI 48.3–83.7%) with Poisson regression and of 73.1% (95% CI 53.3–84.5%) with Cox regression. For HPV45, significant decreases of 67.3% (95% CI 8.8–88.3%) and 69.8% (95% CI 15.2–89.3%) were observed. For HPV18, HPV31, and HPV33, the indirect effect was not statistically significant.

**Conclusions:** Large indirect effects of the bivalent HPV vaccination program were observed for HPV16 and HPV45 infections.

## Background

A persistent infection with a high-risk type of human papillomavirus (HPV) is the established cause of cervical cancer [1]. Since 2006, prophylactic vaccination is available to prevent HPV-associated diseases. High vaccine efficacy and vaccine effectiveness against persisting vaccine-type infections have been observed for licenced HPV vaccines [2-4]. Prolonged high vaccination coverage has led to a reduction in the incidence of HPV vaccine-type infections, precancerous lesions and cervical cancer [5, 6]. In countries with suboptimal HPV vaccination coverage, it is pivotal to assess the magnitude of herd effects in the unvaccinated population.

In the Netherlands, the bivalent HPV (2vHPV) vaccine was introduced in the national immunisation program in 2010, which was preceded by a catch-up campaign in 2009 for girls born between 1993 and 1996. Originally, girls were offered HPV vaccination via a 3-dose (3D) schedule in the year they turned 13. In 2014, the 3D schedule was replaced by a 2-dose (2D) schedule [7-9]. The coverage of the 2vHPV vaccine within the immunisation program has been relatively low for many years, with a coverage ranging between 46% and 63% [10]. In recent years, the program has been expanded to include boys and catch-up campaigns have been temporarily implemented for individuals up to 28 years of age. The low coverage over an extended period has left a significant portion of the population susceptible to vaccine-related HPV type infections. Besides direct vaccine effectiveness, vaccination can offer indirect protection through herd immunity, benefiting everyone in the population, regardless of their vaccination status [11].

The magnitude of herd effects in the unvaccinated population has been studied in only a few studies [12-15]. This is even more true for the 2vHPV vaccine. These studies indicated a reduction in HPV16/18 infections and cervical intraepithelial neoplasia grade 3 (CIN3) rates among unvaccinated women in the postvaccination period than in the prevaccination period. However, these studies did not follow a cohort over time or adjust for sociodemographic characteristics over time. The data analysed in these studies covered a relatively short period of 6-9 years following the introduction of HPV vaccination.

Here, we assessed the indirect effect of the Dutch routine HPV vaccination program on the occurrence of incident HPV infections using longitudinal data collected from the “HPV Amongst Vaccinated and Non-vaccinated Adolescents” (HAVANA, 3D-eligible) study and the HAVANA-2 (2D-eligible) study. We compared the occurrence of incident HPV infections between similarly aged unvaccinated female participants in the HAVANA study from 2010/11 (i.e shortly after the introduction of HPV vaccination in the immunisation program) until 2015/16 and unvaccinated female participants in the HAVANA-2 study from 2017-2022 [16, 17]. The current analysis includes data spanning a significant period after the introduction of HPV vaccination in the Netherlands, enabling the assessment of herd effects through the comparison of these two cohorts.

## Methods

### Study design and procedures

This study compared the incidence of HPV infections among unvaccinated participants in a cohort study initiated in 2009, the year in which routine HPV vaccination for girls was introduced, using follow-up data from 2010/11- 2015/16 to the incidence of HPV infections among unvaccinated participants in a cohort study initiated in 2016 and using follow-up data from 2017-2022 (Supplementary Figure 1). The design of both studies has been described in detail previously [17, 18]. Briefly, two longitudinal cohort studies were established to estimate the vaccine effectiveness of HPV vaccination following the catch-up campaign in 2009/10 (HAVANA, birth cohort 1993/94 eligible for three doses of HPV vaccination) and following the switch towards a 2D schedule in 2014 (HAVANA-2, birth cohort 2001). Vaccination status was acquired through the national vaccination registration system, Praeventis [19]. The participants were asked to complete an annual web-based questionnaire on demographics and sexual behaviour, and to collect a vaginal self-sample (Viba-Brush; Rovers Medical Devices, Oss, the Netherlands). The studies adhered to the tenets of the Declaration of Helsinki and were approved by the Medical Ethics Committee of the Amsterdam University Medical Center (UMC) in the Netherlands (2009/022). Informed consent was obtained from all participants before their inclusion.

The vaginal self-samples were tested for HPV DNA using the SPF10–DEIA-LiPA25 platform (DDL Diagnostics Laboratory, Rijswijk, the Netherlands) [17, 20]. This assay is able to distinguish high-risk HPV types 16, 18, 31, 33, 35, 39, 45, 51, 52, 56, 58, and 59. Additionally, the assay can detect low-risk HPV types 6, 11, 34, 40, 42, 43, 44, 53, 54, 66, 70, and 74.

### Statistical analysis

Unvaccinated participants from both cohort studies were included in the current analyses. A baseline measurement before vaccination was available for the HAVANA cohort, but not for the HAVANA-2 cohort. The baseline measurement from the HAVANA cohort and the first-round measurement from the HAVANA-2 cohort were used to select participants who were HPV negative for all types to ensure that participants were comparable with regard to HPV status for the main analyses. Data from the second up to and including the seventh round (2017-2022) of the HAVANA-2 cohort were compared with data from the first round (2010/11) up to and including the sixth round (2015/16) of the HAVANA cohort. Thus data from unvaccinated women 8-13 years after the introduction of vaccination in 2009 were compared with data from unvaccinated women 1-7 years after introduction. The mean age per study round between the two cohorts was similar.

For each study round, the individual HPV type-specific infection status was retrieved. The first recorded type-specific infection was considered within an individual, and participants were subsequently censored. The time of infection onset was established as the midpoint between the last negative HPV test and the first positive HPV test.

The indirect effect of vaccination was estimated for the combined vaccine types HPV16/18 and cross-protective types HPV31/33/45, as well as for HPV types 16, 18, 31, 33, and 45 separately. The combined endpoint was defined as having an infection with at least one of these HPV types. The combined incidence of non-cross-protective HPV types 35, 39, 51, 52, 56, 58, and 59 was included in the analyses to correct for differences in the background prevalence between the two cohorts (see below).

Two regression methods were used. In the first method, Poisson regression was used to calculate ratios of incidence rates (IRRs) of a HPV vaccine or cross-protective type (or combined HPV types 16/18 and HPV types 31/33/45) in HAVANA-2 versus HAVANA. IRRs were also calculated for non-cross-protective types. Subsequently, the indirect effect was estimated by the ratio of the IRR of a vaccine or cross-protective HPV type (or combined HPV types) and the IRR of non-cross-protective types. Estimates were accompanied by 95% confidence intervals (95% CIs). The indirect effect was defined as (1–ratio IRR) × 100%. In the second method, the Cox proportional hazard (PH) model was used to calculate hazard ratios (HRs) for the incidence of the vaccine or cross-protective HPV type and combinations of HPV types among the participants of both cohorts. The HRs were adjusted for the time-varying sociodemographic characteristics age (continuous), number of lifetime sexual partners (0, 1-3, ≥4), contraception use (yes/no), degree of urbanisation (low/high), and educational level (low/high). The PH assumption was checked for each variable by visually inspecting the log-log plots and the Schoenfeld residual plots. The indirect effect was defined as (1–HR) × 100%. All analyses were performed using SAS (version 9.4). P-values < .05 were considered statistically significant.

### Sensitivity analyses

The analyses were repeated under the assumption that participants were only excluded if they tested positive for (at least one of) the HPV type(s) at baseline. In a second sensitivity analysis, HPV types 35, 52, 58, and 59 were excluded from the non-cross-protective types because the evidence on the lack of cross-protection is ambiguous for types 35, 52, and 58 [21], and because there is some evidence that the SPF10–DEIA-LiPA25 HPV DNA test has a reduced analytical sensitivity for the detection of type 59 [22].

## Results

A total of 1588 unvaccinated participants were included in the analyses, 661 (41.6%) of whom were eligible for a 3D vaccination schedule in 2009 (HAVANA participants) and 927 (58.4%) were eligible for a 2D vaccination schedule in 2014 (HAVANA-2 participants) (Figure 1). The demographic and sexual behavior characteristics of the unvaccinated study participants in both cohorts are summarised in Table 1. At the start of the study period, a greater proportion of HAVANA participants (compared with HAVANA-2 participants) lived in highly urbanised areas (68.0% *vs*. 51.0%), had their sexual debut (42.4% *vs*. 26.6%), were current smokers (30.1% *vs*. 22.1%) and ever used contraception (58.3% *vs*. 44.6%). During follow-up, most of these differences decreased; however, a larger proportion of HAVANA-2 participants achieved a high education level than HAVANA participants did (Table 1, P-value < .0001).

**Figure 1.**
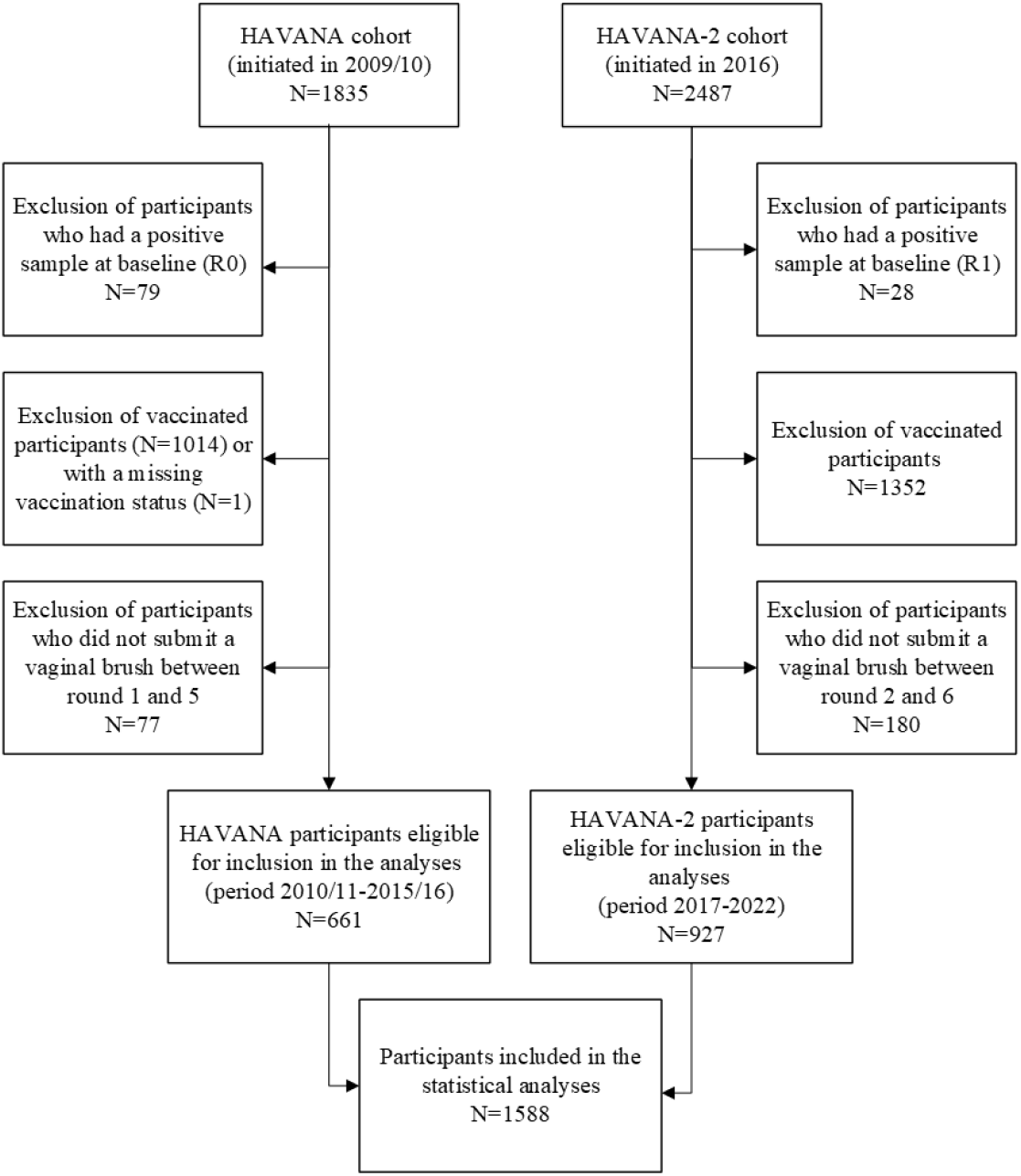
Schematic representation of the exclusion criteria used to construct the population under study

**Table 1.**
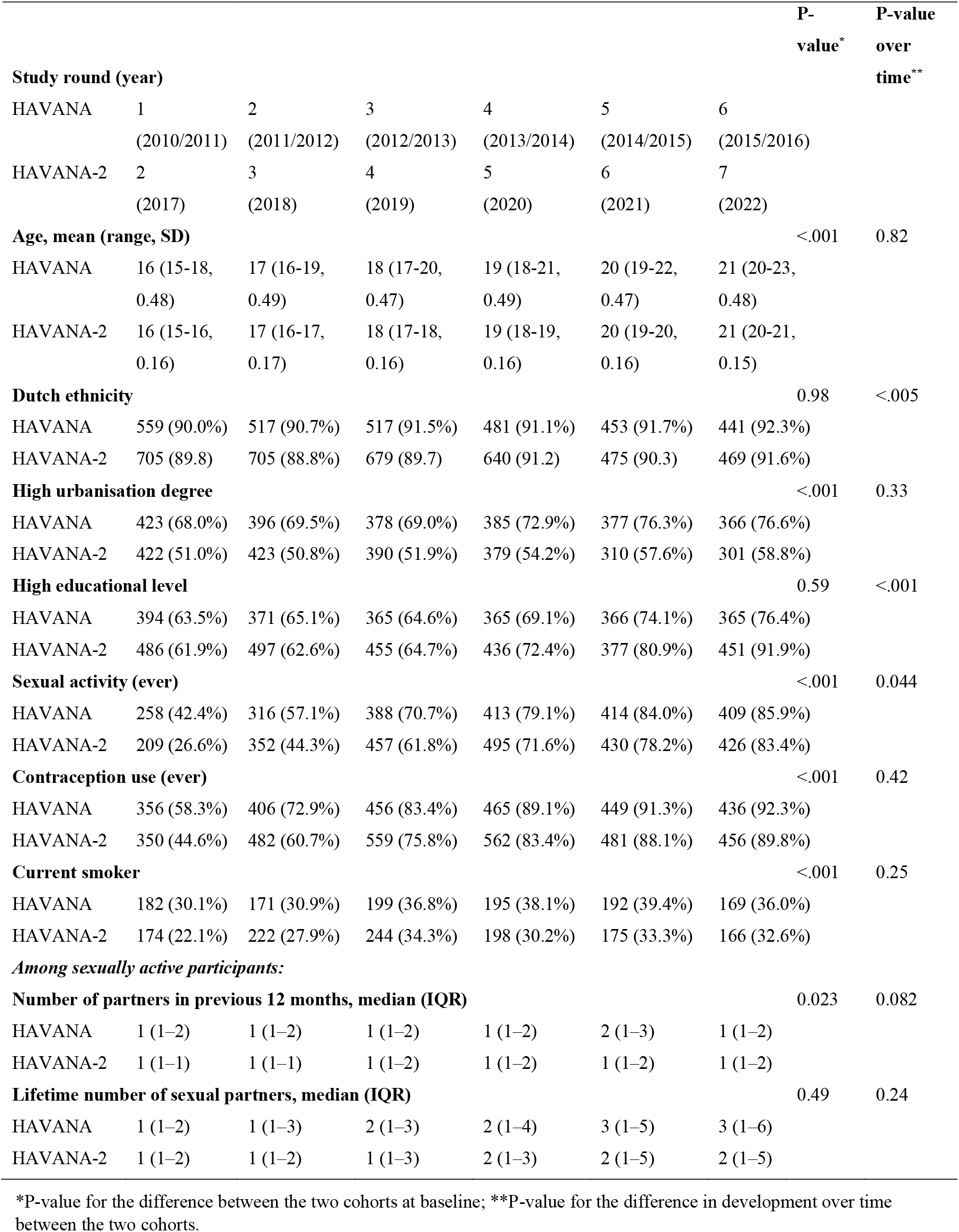
Characteristics of the unvaccinated participants in the HAVANA and HAVANA-2 studies over time

### Poisson regression analyses

The incidence of HPV16 and HPV45 infections, corrected for the incidence of non-cross-protective types, was significantly lower among unvaccinated HAVANA-2 participants than among unvaccinated HAVANA participants, with indirect effects of 70.9% (95% CI 48.3–83.7%) and 67.3% (95% CI 8.8–88.3%), respectively (Table 2). For HPV18, HPV31, and HPV33, the protective indirect effect estimates were 42.4% (95% CI -9.7– 69.7%), 29.3% (95% CI -20.0–58.4%), and 39.4% (95% CI -31.0–72.0%), respectively, and these estimates were not statistically significant. For the estimate of combined HPV types 16/18, an indirect effect of 59.0% (95% CI 35.5–74.0%) was observed. A decreased incidence of HPV31/33/45 infections was also observed, although the decrease was not statistically significant (36.1%, 95% CI -1.1–59.6%).

**Table 2.**
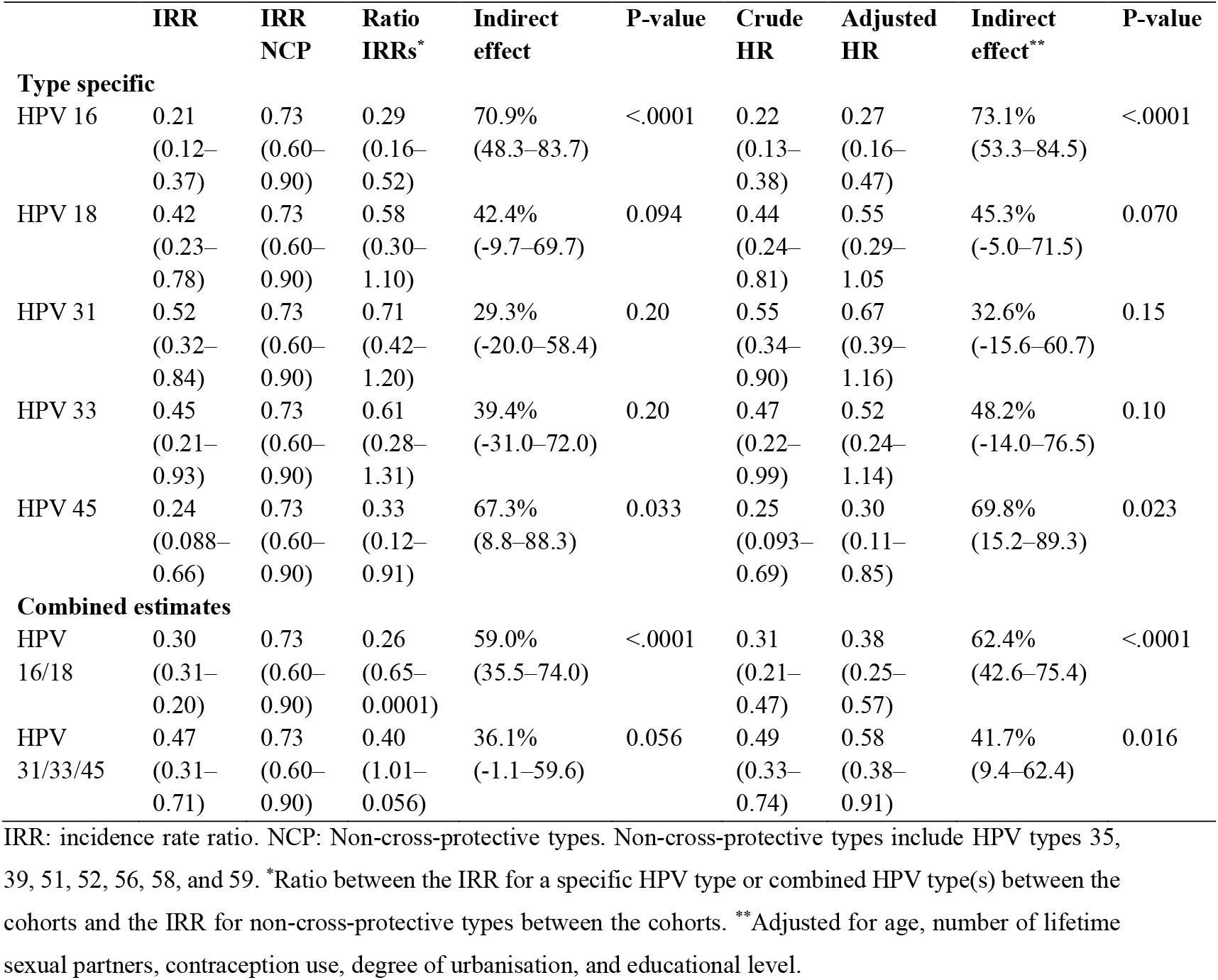
Indirect effect estimates in unvaccinated HAVANA-2 versus HAVANA participants, obtained from Poisson and Cox regression

### Cox regression analyses

Type-specific Kaplan Meier curves are shown in Supplementary Figure 2. The results of the Cox PH analyses comparing HAVANA-2 with HAVANA participants, both at a type-specific level and for the outcomes of combined HPV types, are displayed in Table 2. The Cox analysis yielded similar results for the indirect effects against incident infections as the Poisson analysis and estimates were 73.1% (95% CI 53.3–84.5%) and 69.8% (95% CI 15.2–89.3%) for HPV16 and HPV45 respectively. For HPV 18, 31, and 45, the indirect effects were not significant: 45.3% (95% CI -5.0–71.5%), 32.6% (95% CI -15.6–60.7%), and 48.2% (95% CI -14.0–76.5%), respectively. For the combined outcomes, a significant indirect effect against incident HPV16/18 infections was observed (62.4%, 95% CI 42.6–75.4%), as well as against incident HPV31/33/45 infections (41.7%, 95% CI 9.4– 62.4%).

#### Sensitivity analyses

Overall, the magnitude and direction of the estimated effects for all HPV types in the sensitivity analyses were comparable to those in the main analyses (Supplementary Table 1). The indirect effect of the vaccine against incident HPV16 and HPV45 infections remained significant in all sensitivity analyses. The exclusion of participants who tested positive for the particular HPV type(s) under study at baseline (rather than the exclusion of participants who tested positive for any type) resulted in slightly higher estimates in the Poisson regression analysis than in the main Poisson regression analysis. In contrast, this type of exclusion in the Cox regression analysis slightly decreased the estimates compared with the estimates from the main Cox regression analysis (Table S1).

## Discussion

In this observational cohort study, we assessed the indirect effect of 2vHPV vaccination by comparing cohort data 8–13 years after the introduction of the HPV vaccine in the Dutch immunisation program with cohort data 1–7 years after introduction. Our evaluation involved a comparison between unvaccinated participants from the HAVANA-2 cohort (birth cohort 2001, 2D vaccination implementation in 2014, data collected in 2017–2022) and unvaccinated participants from the HAVANA cohort (birth cohorts 1993/94, 3D vaccination implementation in 2009, data collected in 2010/11–2015/16). The outcomes of two types of regression analyses revealed a reduction of approximately 70% in incident HPV16 infections in the HAVANA-2 study compared with the HAVANA study, despite the relatively low vaccination coverage in the Netherlands [23]. An indirect effect of around 70% against incident HPV45 infections was also found. The outcomes further suggest a reduction in incident HPV types 18, 31, and 33, although statistical significance was not reached.

To our knowledge, there have been limited publications on the long-term indirect effects of 2vHPV vaccination programs on the occurrence of infections. Previous studies from Scotland and Spain that assessed the indirect effect of vaccination on HPV infections were repeated cross-sectional studies, whereas we followed individual women from two cohorts over time which increases the validity of our study. Previous studies used data collected over a shorter period (up to 9 years) after the introduction of HPV vaccination. These studies reported a 30–33% reduction in the prevalence of HPV16/18 infections among women aged 16–26 years who were attending a health care centre or participating in a cervical screening program [12, 13]. The greater reduction rates of HPV16 and HPV types 16 and 18 combined, as observed in our study, might be attributable to the longer follow-up period of our study. In line with the findings in our study, Markowitz et al. reported a decrease in the prevalence of HPV16/18 of approximately 53% among unvaccinated women aged 20–24 years who participated in cervical cancer screening up to 10 years after the introduction of the quadrivalent HPV vaccine [14]. The non-significance of the indirect effect estimates that we observed for cross-protective HPV types 31 and 33 infections might be related to the sample size of the cohorts. However, the point estimates indicated that the indirect effect of the cross-protective types may still yield a substantial impact at the population level. In combination with the direct protection estimates against cross-protective types infections reported by Hoes et al. [16, 17], we assume a substantial total effect on the general population.

This study used two different regression methods to quantify the indirect impact of the vaccination program in the Netherlands. In our first method, we corrected for changes over time in the type-specific incidence of HPV by assuming that changes in the non-cross-protective HPV types reflect changes in the background HPV infection risk. This type of approach was recently suggested by Sasieni [24]. We also used a Cox PH model approach, which allows for adjustments for time-varying covariates when estimating vaccine effects. The two different regression methods gave highly comparable results with regard to the magnitude and direction of the estimates for most HPV types. The close agreement between the models indicates the robustness of our findings regarding indirect protection. We performed three sensitivity analyses to verify whether data classification choices would substantially affect the estimated outcomes. Overall, we observed similar results between the outcomes of the main analyses and the three sensitivity analyses, which underlines the robustness of the estimates. The significant indirect effects we observed in this study are pivotal as they contribute to equalising the cervical cancer risk of vaccinated and unvaccinated women when they participate in a cervical cancer screening program, hence, facilitating the integration of vaccination and screening.

A key strength of our study is the use of observational data from two longitudinal cohort studies. The use of data covering the period 2009 to 2022 ensured sufficient statistical power to show declines in incident HPV16 infections despite the relatively low vaccine coverage in the Netherlands. The use of longitudinal data retrieved from annual questionnaires allowed us to control for time-varying variables in the Cox PH regression analyses. Furthermore, the data allowed type-specific exclusion at baseline to analyse that particular HPV type. Another strength of our study is that we used different regression methods, which demonstrated nearly similar results. Nevertheless, certain limitations should be acknowledged. A baseline measurement before vaccination was not available for the HAVANA-2 cohort, which we mitigated by using the first study year as a baseline measurement. In both cohorts, the educational level was slightly higher and participants were less likely to be non-Dutch compared to the general Dutch population, which might affect the generalizability of the findings [25, 26]. However, more than half of the population had their sexual debut at 18 years of age, which is similar to the median age of sexual debut among girls in the general Dutch population [27]. Finally, reliance on self-reported data for sociodemographics and risk behaviour potentially affects validity.

## Conclusions

To conclude, we observed a high indirect effect of the 2vHPV vaccine against HPV16 and HPV45 infections among unvaccinated girls 8–13 years after the introduction of HPV vaccination into the Dutch immunisation program, thereby contributing to the success of HPV vaccination programs. The extent to which unvaccinated women are protected against vaccine-related HPV infections is expected to increase in the future as the Netherlands recently (2022) switched to gender-neutral HPV vaccination. The indirect effect of vaccination observed in our study, combined with the earlier demonstrated direct vaccine effectiveness, indicates a considerable total effect on the general population. Continued surveillance of both individual- and population-level effects of the HPV vaccination program is crucial to monitor future changes in protection or shifts in HPV trends over time and could be important for guiding cervical cancer screening recommendations.

## Data Availability

Aggregated data that support the findings of this study are available upon reasonable request from the corresponding author.

## List of abbreviations

HPV: human papillomavirus
IRR: incidence rate ratio
HR: hazard ratio
2vHPV: bivalent HPV
3D: 3-dose
2D: 2-dose
CIN3: cervical intraepithelial neoplasia grade 3
HAVANA study: HPV Amongst Vaccinated and Non-vaccinated Adolescents study
CI: confidence intervals
PH: proportional hazard
HR: hazard ratio

## Declarations

### Ethics approval and consent to participate

The studies were approved by the Medical Ethics Committee of the Amsterdam University Medical Center (UMC), the Netherlands (2009/022). All participants provided written informed consent before inclusion.

### Consent for publication

Not applicable.

### Availability of data and materials

The statistical datasets analysed during the current study are available from the corresponding author on reasonable request and with permission of National Institute for Public Health and the Environment.

### Authors’ contributions

HEdM, AK, and MM were involved in the original study design. MM and JD were involved in the data collection. AK was responsible for the laboratory analyses. MM and JD performed the statistical analysis, with contributions from MK, JB, and HEdM. MM prepared the first draft of the manuscript. All authors contributed to the data interpretation and revision of the manuscript before submission, and approved the final version of the manuscript.

## Acknowledgements

We would like to thank Najima, Tsira, Thomas, Naomi, Suzan and Elske (Centre for Infectious Disease Research, Diagnostics and laboratory Surveillance [IDS], National Institute for Public Health and the Environment, the Netherlands) for their contribution to the laboratory analysis of the samples.

## References

1. Walboomers JM, Jacobs MV, Manos MM, Bosch FX, Kummer JA, Shah KV, et al. Human papillomavirus is a necessary cause of invasive cervical cancer worldwide. The Journal of pathology. 1999;189(1):12–9.

2. Markowitz LE, Drolet M, Perez N, Jit M, Brisson M. Human papillomavirus vaccine effectiveness by number of doses: Systematic review of data from national immunization programs. Vaccine. 2018;36(32, Part A):4806–15.

3. Lehtinen M, Paavonen J, Wheeler CM, Jaisamrarn U, Garland SM, Castellsagué X, et al. Overall efficacy of HPV-16/18 AS04-adjuvanted vaccine against grade 3 or greater cervical intraepithelial neoplasia: 4-year end-of-study analysis of the randomised, double-blind PATRICIA trial. The lancet oncology. 2012;13(1):89–99.

4. Hildesheim A, Wacholder S, Catteau G, Struyf F, Dubin G, Herrero R, et al. Efficacy of the HPV-16/18 vaccine: final according to protocol results from the blinded phase of the randomized Costa Rica HPV-16/18 vaccine trial. Vaccine. 2014;32(39):5087–97.

5. Rahangdale L, Mungo C, O’Connor S, Chibwesha CJ, Brewer NT. Human papillomavirus vaccination and cervical cancer risk. bmj. 2022;379.

6. Kavanagh K, Pollock K, Potts A, Love J, Cuschieri K, Cubie H, et al. Introduction and sustained high coverage of the HPV bivalent vaccine leads to a reduction in prevalence of HPV 16/18 and closely related HPV types. British journal of cancer. 2014;110(11):2804–11.

7. National Institute for Public Health and the Environment, (RIVM). Wijziging HPV-vaccinatieschema: 1 prik minder. Infectieziekten bulletin. 2014;25.

8. European Medicines Agency (EMA). Assessment report Cervarix, procedure number EMEA/H/C/000721/II/0048. 2013.

9. European Medicines Agency (EMA). Assessment report Cervarix, procedure number EMEA/H/C/000721/II/0036. 2013.

10. Pluijmaekers AJM, de Melker HE. The National Immunisation Programme in the Netherlands. Surveillance and developments in 2022-2023. 2023.

11. S H, K B, P F. Vaccination Programmes: Epidemiology, Monitoring, Evaluation. 1st edition ed. London: Routledge; 2021.

12. Cameron RL, Kavanagh K, Pan J, Love J, Cuschieri K, Robertson C, et al. Human papillomavirus prevalence and herd immunity after introduction of vaccination program, Scotland, 2009–2013. Emerging infectious diseases. 2016;22(1):56.

13. Purriños-Hermida MJ, Santiago-Pérez MI, Treviño M, Dopazo R, Cañizares A, Bonacho I, et al. Direct, indirect and total effectiveness of bivalent HPV vaccine in women in Galicia, Spain. PLoS One. 2018;13(8):e0201653.

14. Markowitz LE, Naleway AL, Lewis RM, Crane B, Querec TD, Weinmann S, et al. Declines in HPV vaccine type prevalence in women screened for cervical cancer in the United States: Evidence of direct and herd effects of vaccination. Vaccine. 2019;37(29):3918–24.

15. Tabrizi SN, Brotherton JM, Kaldor JM, Skinner SR, Liu B, Bateson D, et al. Assessment of herd immunity and cross-protection after a human papillomavirus vaccination programme in Australia: a repeat cross-sectional study. The Lancet infectious diseases. 2014;14(10):958–66.

16. Hoes J, King AJ, Berkhof J, de Melker HE. High vaccine effectiveness persists for ten years after HPV16/18 vaccination among young Dutch women. Vaccine. 2023;41(2):285–9.

17. Hoes J, King AJ, Klooster TMSvt, Berkhof J, Bogaards JA, de Melker HE. Vaccine Effectiveness Following Routine Immunization With Bivalent Human Papillomavirus (HPV) Vaccine: Protection Against Incident Genital HPV Infections From a Reduced-Dosing Schedule. The Journal of infectious diseases. 2022;226(4):634–43.

18. Donken R, King A, Bogaards J, Woestenberg P, Meijer C, De Melker H. High effectiveness of the bivalent human papillomavirus (HPV) vaccine against incident and persistent HPV infections up to 6 years after vaccination in young Dutch women. The Journal of Infectious Diseases. 2018;217(10):1579–89.

19. van Lier A, Oomen P, de Hoogh P, Drijfhout I, Elsinghorst B, Kemmeren J, et al. Praeventis, the immunisation register of the Netherlands: a tool to evaluate the National Immunisation Programme. Eurosurveillance. 2012;17(17):20153.

20. van Eer K, Middeldorp M, Dzebisasjvili T, Lamkaraf N, de Melker HE, Steenbergen RD, et al. Effects of 2 and 3 Vaccinations With the Bivalent Human Papillomavirus (HPV) Vaccine on the Prevalence and Load of HPV in Clearing and Persistent Infections in Young Women. The Journal of Infectious Diseases. 2023:jiad080.

21. Brown DR, Joura EA, Yen GP, Kothari S, Luxembourg A, Saah A, et al. Systematic literature review of cross-protective effect of HPV vaccines based on data from randomized clinical trials and real-world evidence. Vaccine. 2021;39(16):2224–36.

22. van Eer K, Leussink S, Severs TT, Marm-Wattimena Nv, Woestenberg PJ, Bogaards JA, et al. Evidence for Missing Positive Results for Human Papilloma Virus 45 (HPV-45) and HPV-59 with the SPF10-DEIA-LiPA25 (Version 1) Platform Compared to Type-Specific Real-Time Quantitative PCR Assays and Impact on Vaccine Effectiveness Estimates. Journal of Clinical Microbiology. 2020;58(11):10.1128/jcm.01626-20.

23. van Lier A, Hament J-M, Knijff M, Westra M, Ernst A, Giesbers H, et al. Vaccination coverage and annual report of the National Immunisation Programme in the Netherlands, 2022. 2023. Contract No.: 2023-0031.

24. Sasieni P. Alternative analysis of the data from a HPV vaccine study in India. The Lancet Oncology. 2022;23(1):e9.

25. Statistics Netherlands, (CBS). Dashboard Population, Origin. Available from: https://www.cbs.nl/en-gb/visualisations/dashboard-population/origin. Accessed on 30 April 2024.

26. Statistics Netherlands, (CBS). Educational level 16+ according to age and sex, 2022 2023. Available from: https://www.cbs.nl/nl-nl/maatwerk/2023/44/opleidingsniveau-16-naar-leeftijd-en-geslacht-2022. Accessed on 30 April 2024.

27. de Graaf H, Vermey K, Oldenhof A, Kraan Y, Beek T, Kuiper L. Seks onder je 25e: Rutgers/Soa Aids Nederland; 2023. Available from: https://rutgers.nl/onderzoeken/seks-onder-je-25e-2023/. Accessed on 2 Juni 2024.

