## Supplementary material for "The indirect effect of the bivalent human papillomavirus vaccination program: an observational cohort study"

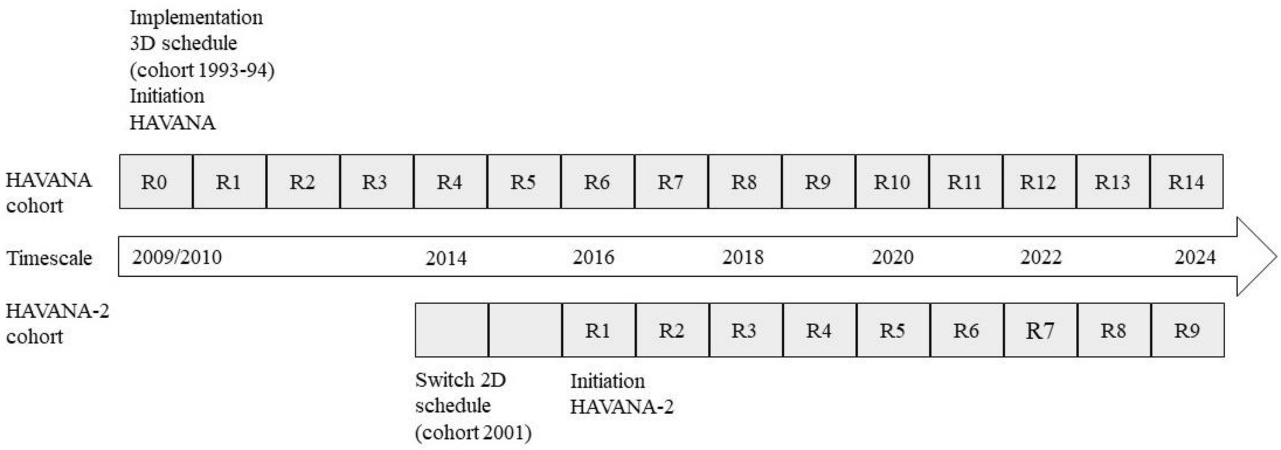


**Supplementary Figure 1.** Schematic representation of the study design of the HAVANA(2) cohorts

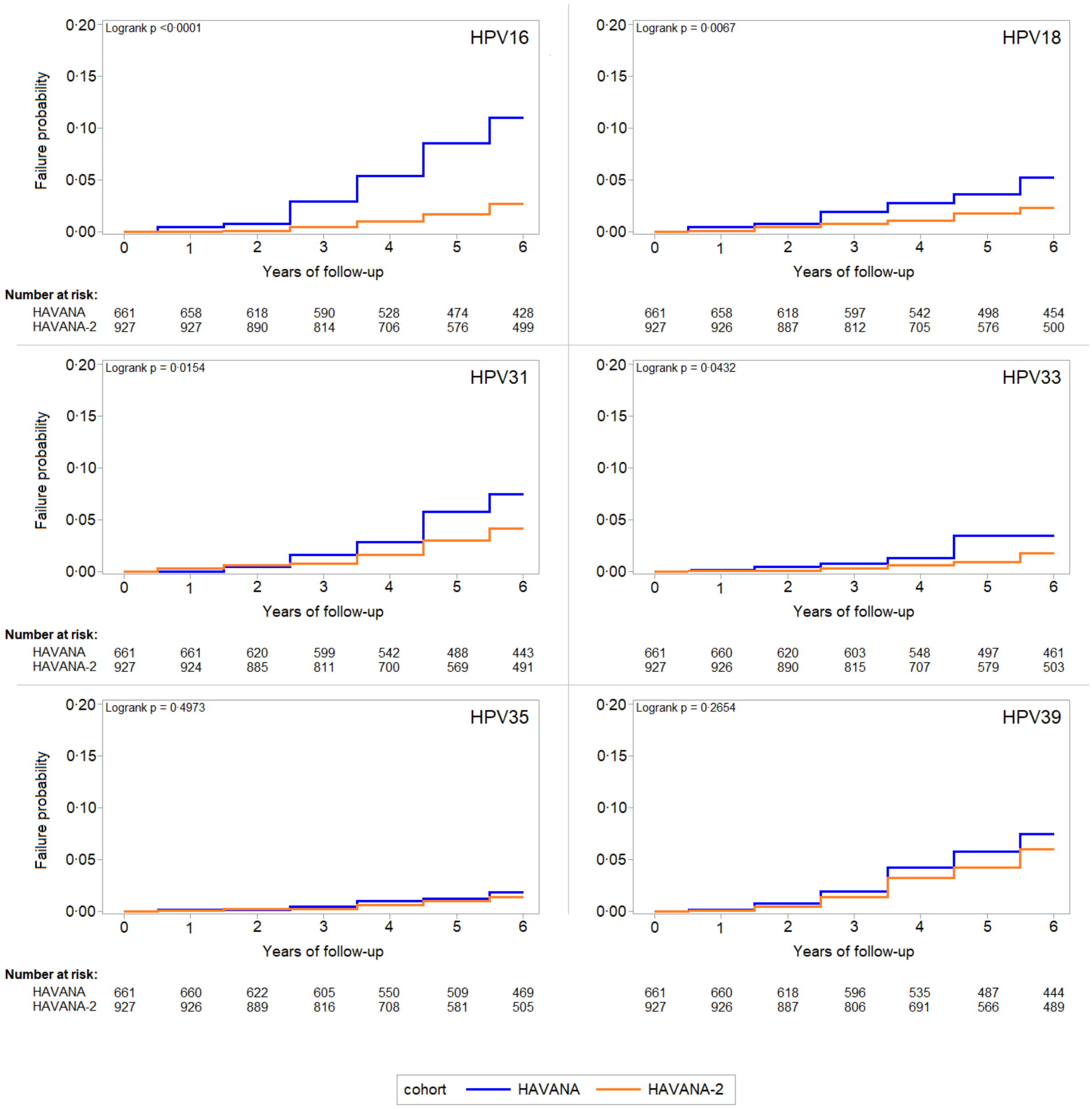
HPV16
Logrank p <0·0001
HPV18
Logrank p = 0·0067
HPV31
Logrank p = 0·0154
HPV33
Logrank p = 0·0432
HPV35
Logrank p = 0·4973
HPV39
Logrank p = 0·2654
Failure probability
0·00 0·05 0·10 0·15 0·20
Years of follow-up
0 1 2 3 4 5 6
Number at risk:
HAVANA 661 658 618 590 528 474 428
HAVANA-2 927 927 890 814 706 576 499
661 658 618 597 542 498 454
927 926 887 812 705 576 500
HAVANA 661 661 620 599 542 488 443
HAVANA-2 927 924 885 811 700 569 491
661 660 620 603 548 497 461
927 926 890 815 707 579 503
HAVANA 661 660 622 605 550 509 469
HAVANA-2 927 926 889 816 708 581 505
661 660 618 596 535 487 444
927 926 887 806 691 566 489
cohort
HAVANA
HAVANA-2

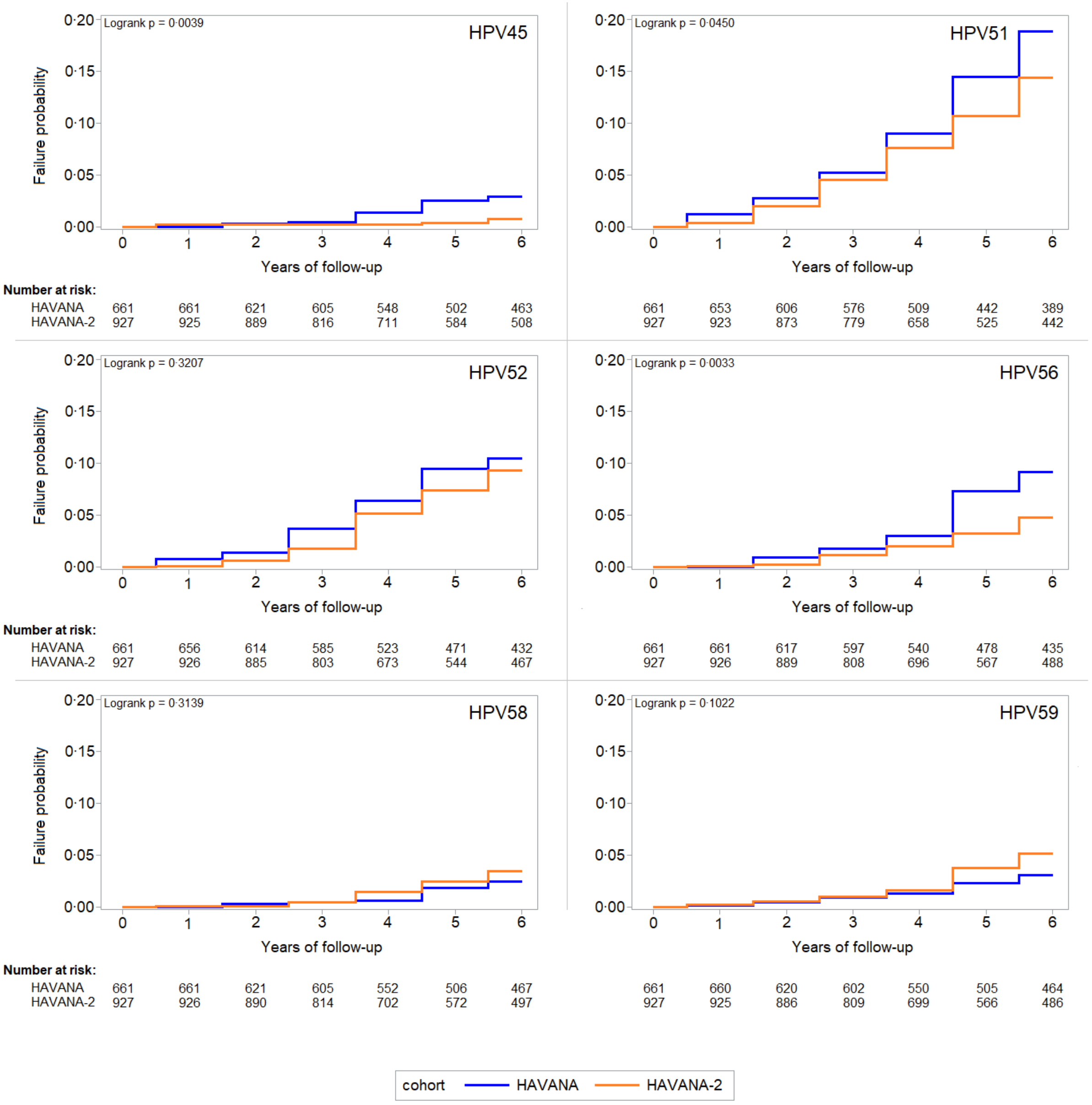


**Supplementary Figure 2.** Kaplan Meier curves of type-specific HPV infections

**Supplementary Table 1.** Results from sensitivity analyses as obtained from the Poisson regression analyses and Cox regression analyses

| | Indirect effect* | P-value | Indirect effect** | P-value | Adjusted indirect effect*** | P-value |
|---|---|---|---|---|---|---|
| **Type specific** | | | | | | |
| HPV 16 | 68.4% (43.2–82.4%) | <.0001 | 71.7% (50.6–83.8%) | <.0001 | 68.4% (45.3–81.7%) | <.0001 |
| HPV 18 | 37.3% (-20.4–67.4%) | 0.16 | 45.0% (-0.5–69.9%) | 0.052 | 44.2% (-5.0–70.3%) | 0.071 |
| HPV 31 | 32.2% (-31.9–55.2%) | 0.34 | 32.4% (-12.1–59.3%) | 0.13 | 28.4% (-23.0–58.4%) | 0.23 |
| HPV 33 | 34.1% (-43.5–69.7%) | 0.29 | 43.5% (-20.4–73.5%) | 0.14 | 42.2% (-28.2–73.9%) | 0.18 |
| HPV 45 | 64.4% (0.2–87.3%) | 0.045 | 67.7% (17.9–87.3%) | 0.018 | 64.8% (6.1-86–8%) | 0.037 |
| **Estimates of combined HPV types** | | | | | | |
| HPV 16/18 | 55.4% (29.0–72.0%) | 0.0007 | 60.0% (38.0–74.2%) | <.0001 | 55.9% (33.0–71.0%) | <.0001 |
| HPV 31/33/45 | 30.5% (-11.3–56.6%) | 0.13 | 38.3% (3.8–60.4%) | 0.033 | 35.1% (-1.2–58.4%) | 0.056 |

* Poisson regression analyses where HPV39, HPV51, and HPV56 were considered non-cross-protective;
** Poisson regression analyses where only participants who tested positive for a particular HPV type were excluded in that type-specific analysis;
*** Cox regression analyses where participants who tested positive for a particular HPV type were excluded in that type-specific analysis.
